# Cascaded Multimodal Deep Learning in the Differential Diagnosis, Progression Prediction, and Staging of Alzheimer’s and Frontotemporal Dementia

**DOI:** 10.1101/2024.09.23.24314186

**Authors:** Gianmarco Guarnier, Janis Reinelt, Eóin N. Molloy, Paul Glad Mihai, Pegah Einaliyan, Sofie Valk, Augusta Modestino, Matteo Ugolini, Karsten Mueller, Qiong Wu, Anahit Babayan, Marco Castellaro, Arno Villringer, Nico Scherf, Konstantin Thierbach, Matthias L. Schroeter, the Alzheimer’s Disease Neuroimaging Initiative, the Australian Imaging Biomarkers and Lifestyle flagship study of ageing, the Frontotemporal Lobar Degeneration Neuroimaging Initiative

## Abstract

Dementia is a complex condition whose multifaceted nature poses significant challenges in the diagnosis, prognosis, and treatment of patients. Despite the availability of large open-source data fueling a wealth of promising research, effective translation of preclinical findings to clinical practice remains difficult. This barrier is largely due to the complexity of unstructured and disparate preclinical and clinical data, which traditional analytical methods struggle to handle. Novel analytical techniques involving Deep Learning (DL), however, are gaining significant traction in this regard. Here, we have investigated the potential of a cascaded multimodal DL-based system (TelDem), assessing the ability to integrate and analyze a large, heterogeneous dataset (n=7,159 patients), applied to three clinically relevant use cases. Using a Cascaded Multi-Modal Mixing Transformer (CMT), we assessed TelDem’s validity and (using a Cross-Modal Fusion Norm - CMFN) model explainability in (i) differential diagnosis between healthy individuals, AD, and three sub-types of frontotemporal lobar degeneration (ii) disease staging from healthy cognition to mild cognitive impairment (MCI) and AD, and (iii) predicting progression from MCI to AD. Our findings show that the CMT enhances diagnostic and prognostic accuracy when incorporating multimodal data compared to unimodal modeling and that cerebrospinal fluid (CSF) biomarkers play a key role in accurate model decision making. These results reinforce the power of DL technology in tapping deeper into already existing data, thereby accelerating preclinical dementia research by utilizing clinically relevant information to disentangle complex dementia pathophysiology.

## Introduction

Dementia significantly reduces patient quality of life and represents a major source of economic and societal burden worldwide^1^. Despite an expected increase in the prevalence of dementia over the coming decades^2^, accurate diagnosis, prognosis, and identification of novel treatment avenues continue to pose significant challenges. After decades of clinical trial setbacks, the emergence of the first disease-modifying therapies (DMTs) targeting amyloid accumulation for Alzheimer’s Disease (AD - which represents a significant proportion of people with dementia^3^) have been reported^4–8^. However, challenges persist as these DMTs show only modest delays in disease progression, need real-world replication of clinical effects, and have a high prevalence of adverse effects^9^. While the roles of amyloid and tau in AD pathophysiology are widely recognized^10^ evidence suggests that the disease extends far beyond a simplistic interaction of these two proteins^11,12^. This multifactorial nature, which involves inflammation^13^, lifestyle factors^14,15^, and APOE ε4 genotype and its complex interactions with brain function^16,17^ has contributed to the high rate of trial failure^18^ and the occurrence of significant adverse reactions to therapy^19,20^. As a result, the efficacy of these therapies remains controversial.

In this evolving landscape, quantifiable biomarkers have emerged as an indispensable resource for AD and dementia research^21^, offering insights into disease progression, differential diagnosis, and therapeutic response^22^. Nevertheless, a noticeable disparity exists between clinical endpoints such as cognitive decline and the preclinical indicators of these biomarkers. This disparity underscores the challenges of translating actionable biomarkers into tangible clinical benefits^23^. However, with more comprehensive and diverse data, new and more precise biomarkers can be identified. This, in turn, could allow for the development of more effective therapeutic targets and more accurate predictors of clinical outcomes. Recent decades have seen a proliferation of large-scale initiatives aimed at achieving this goal through datasets such as the Alzheimer’s Disease Neuroimaging Initiative (ADNI) and its various incarnations^24^, including the Australian Imaging Biomarkers and Lifestyle Study (AIBL)^25^. These and similar large multimodal datasets contain a wealth of clinically relevant information that, if harnessed effectively, could revolutionize dementia research, enhance clinical trial design and execution, and ultimately improve standard of care and patient outcomes.

The full potential of these datasets, however, remains somewhat unexploited. This is due to challenges in analyzing data obtained from numerous sources, each having unique issues such as data-missingness and comparability^26,27^. Standard analytical techniques, although sophisticated and well-validated, are often ill-equipped to handle this lack of harmonization and structure, particularly the multimodality ^28^ associated with large datasets. This absence of uniformity thus undermines the fruitful translation of preclinical research to clinical application. Recent years, however, have seen the advent of new technologies that could address these barriers. The rapidly advancing field of Deep Learning (DL) for example, presents such an opportunity and is gaining significant traction within regulatory frameworks in North America^29^, Europe^30^, and Asia^31^. Recent innovations, particularly in multimodal DL models, offer an unprecedented opportunity for more holistic analyzes of these heterogeneous dementia datasets^32^, even in the presence of missing and diverse forms of data^33^ As a result, DL-powered analytical technologies could enhance how we approach, understand, and analyze large, complex, and disorganized data.

While promising in this respect, these multimodal DL systems require diligent validation due to (i) the often-subtle nature of novel biomarker identification, (ii) the need for alignment of model predictions with existing medical knowledge, and (iii) the “black box” nature of model decision-making. In this paper, we aimed to provide this validation of a DL-powered clinical decision support system designed to enhance dementia research and patient care. Termed the TelDem system, we evaluated the power of this DL approach in integrating multimodal data from disparate sources, handling missing data, and incorporating diverse modalities. Specifically, we focused on six datasets from the ADNI, AIBL and the Frontotemporal Lobar Degeneration Neuroimaging Initiative (FTLDNI) studies, comprising of over 7,000 patients with AD and three subtypes of frontotemporal lobar degeneration (FTLD). Additionally, we defined three distinct use cases, each reflecting real-world clinical scenarios based on expert opinion. In the first use case (UC1), we assessed TelDem in the context of the often-difficult differential diagnosis of AD and FTLD^34,35^ by evaluating the architecture’s ability to successfully identify cognitively normal individuals (CN), AD, and three FTLD subclasses (behavioral variant frontotemporal dementia – bvFTD, semantic variant Primary Progressive Aphasia – svPPA, and non-fluent agrammatic variant Primary Progressive Aphasia – nfvPPA). In the second use case (UC2), we focused on AD and its stages including risk-states, evaluating whether the architecture could successfully classify participants into CN, mild cognitive impairment (MCI), or AD. Finally, in the third use case (UC3), we evaluated the system in predicting the conversion of MCI to AD. Each use case was designed to reflect current challenges facing biomarker identification, with the aim of testing TelDem’s potential in accelerating dementia research and patient care.

## Methods

Data used in this article were obtained from six open-source datasets; the Alzheimer’s Disease Neuroimaging Initiative (ADNI)^36^, releases 1, 2, 3, and ADNI-GO, the Australian Imaging, Biomarker & Lifestyle Flagship Study of Ageing (AIBL)^25,37^, and the Frontotemporal Lobar Degeneration Neuroimaging Initiative – (FTLDNI). Each dataset contains a variation of cognitively normal individuals (CN), people with MCI, AD, and FTLD, resulting in 7,159 participants included in our study (Table 1).

**Table 1:** Diagnosis Composition of the harmonized dataset data-points accumulated from three open-source datasets; ADNI, AIBL, and FTLDNI. CN = Cognitively normal individuals, sMCI = Stable mild cognitive impairment, pMCI = progressive Mild Cognitive Impairment, AD = Alzheimer’s disease, bvFTD = behavioral variant Frontotemporal Dementia, svPPA = semantic variant Primary Progressive Aphasia, nfvPPA = non-fluent variant Primary Progressive Aphasia. Other refers to data which did not satisfy our modeling criteria (see Data Selection paragraph).

|  | CN | sMCI | pMCI | AD | bvFTD | svPPA | nvPPA | Other |
| --- | --- | --- | --- | --- | --- | --- | --- | --- |
| **ADNI** | 3067 | 2574 | 966 | 1773 | - | - | - | 720 |
| **AIBL** | 1263 | 193 | 64 | 158 | - | - | - | 8 |
| **FTLDNI** | 451 | - | - | - | 264 | 147 | 137 | 9 |

ADNI (https://adni.loni.usc.edu/) was launched in 2003 as a public-private partnership, led by Principal Investigator Michael W. Weiner, MD. The primary goal of ADNI has been to test whether serial magnetic resonance imaging (MRI), positron emission tomography (PET), other biological markers, and clinical and neuropsychological assessment can be combined to measure the progression of mild cognitive impairment (MCI) and early Alzheimer’s disease (AD). ADNI is a longitudinal multicenter neuroimaging study consisting of CN individuals, MCI, and AD across four separate studies: ADNI-1, ADNI-GO, ADNI-2, and ADNI-3. Participants underwent physical and neurological examinations, standardized neuropsychological tests, and provided blood, urine, and in a minority of cases (∼20%), cerebrospinal fluid (CSF) samples. All participants underwent either a 1.5T or 3T MRI scan, a Mini-Mental State Examination (MMSE), the Clinical Dementia Rating (CDR), and memory performance measured on the Wechsler Memory Scale (WMS-R) Logical Memory (LM-II subscale of the WMS-R) at screening. Eligibility criteria relied on a set of cognitive scores and included, for instance, an MMSE score between 24 and 30 in cognitively normal individuals and MCI, and between 20 and 26 in AD^36^.

AIBL is a two-site longitudinal study by the AIBL study group, in which participants were initially classified as AD, MCI, and CN and assessed over approximately 10 years. Participants were at least 60 years of age and were screened for neurological conditions including non-AD dementia and psychiatric illnesses including schizophrenia, depression, Parkinson’s disease, or stroke. AIBL study methodology has been reported previously^37^.

The FTLDNI study investigates sporadic and familial forms of FTLD, in particular bvFTD, svPPA, and nfvPPA. This includes both symptomatic individuals and asymptomatic or ambiguously symptomatic family members. The study aims to delineate brain function changes attributable to these disorders and differentiate them from normal aging. Participants were categorized after a comprehensive series of physical exams, cognitive evaluations, and interviews according to international consensus criteria^38,39^. Notably, FTLDNI includes two extra CDR readouts that pertain to a modified version of the assessment: the CDR Dementia Staging Instrument PLUS National Alzheimer’s Coordinating Center (NACC) Behavior and Language Domains (CDR plus NACC FTLD^40^). This assessment was specifically aimed at discriminating FTLD from AD. For a detailed breakdown of the compiled dataset endpoints, including full sample demographics, see supplementary table 1.

### Clinical Use Cases

We identified three clinical settings that mirror realistic use cases (UCs) based on clinical expert opinion. These UCs were evaluated by iteratively increasing the number of input modalities, thereby gradually transitioning from a unimodal to a multimodal model. We designed the unimodal application to simulate a real-world situation in which data are limited, inputting only MRI-data to the model. We then expanded to data that are non-invasive and accessible in most clinical environments. This clinical standard setting includes the patient’s demographics and behavioral and cognitive assessments, such as the MMSE. Finally, we evaluated the system in the most comprehensive UC (referred to hereafter as the Invasive/Research setting) where we included biomarkers or variables not acquired routinely. These included CSF biomarkers, plasma biomarkers, and APOE genotyping. In UC1, we tested the system’s ability to distinguish between CN, AD, and the FTLD subclasses – bvFTD, svPPA, and nfvPPA). For UC1, we tested the system fits in an unimodal configuration with only T1-w MRI as an input, and subsequently in a multimodal configuration in which demographics, cognitive assessments, and APOE status were added. Secondly, UC2 assessed whether the system could stage AD disease, first in CN participants, MCI, and AD, and, in UC3, the progression from MCI to AD. For each UC, we conducted 5-fold cross-validation experiments by splitting the folds on a subject level to avoid data leakage when evaluating our models (see supplementary Fig. 1).

### Data Selection

We applied quality control on the assigned labels in the ADNI and AIBL datasets by removing participants who had at least one diagnosis not related to AD. Notably, this step allowed us to exclude many MCI participants who were indicated to have FTLD or Lewy Body Dementia, which have very different features from MCI and AD (these participants appear as “Other” in Table 1, alongside people without a diagnosis and conditions outside of our scope).

### Use Case 1 - Dementia Differential Diagnosis

Data selected for this task includes CN participants from all studies, AD patients from AIBL and ADNI, and FTLD subclasses from the FTLDNI dataset. We modelled structural MRI, demographics, behavioral assessments, and cognitive scores as modalities. Among these, we included the CDR, ensuring the score formulation was consistent across the FTLDNI Dataset, and ADNI/AIBL (see supplementary Fig. 2).

### Use Case 2 - AD Staging

Dementia patients came from the AIBL and the ADNI studies while the CN group also included patients from FTLDNI. Modalities included were demographic, behavioral assessments, CSF and plasma biomarkers, and T1-weighted (T1w) Magnetization-Prepared Rapid Gradient-Echo (MPRAGE) MRI. We did not include any cognitive scores since they were used as screening tools and outcome metrics to define the conditions at baseline.

### Use Case 3 - MCI conversion

Only ADNI and AIBL participants were included for this UC. We defined as progressive MCI (pMCI) all MCI patients who also received an AD diagnosis during the duration of the study, independently of the progression time. Data used for this application was demographics, T1w MRI, CSF and plasma biomarkers, behavioral scores, and cognitive scores since their adoption to define the condition only applied to the baseline encounter.

### Preprocessing of Tabular Data

#### Cerebrospinal Fluid and Plasma Biomarkers

We obtained all data from the Imaging and Data Archive (IDA) from the Laboratory of Neuroimaging (LONI) portal and data processing is as described by ADNI. Briefly, CSF Gap_43_ and Neurofilament Light Chain (NFL) were preprocessed with an ELISA assay while plasma NFL was processed with a single molecule array (Simoa) technique. Plasma phosphorylated tau 181 (p-Tau_181_) was assessed with an in-house Simoa array while plasma NT1-Tau was processed through the Quanterix Simoa Platform HD-1. CSF Amyloid β_1-40_ (Aβ_1-40_), Amyloid β_1-42_ (Aβ_1-42_), Tau, and p-Tau were processed by the University of Pennsylvania. We removed outliers by filtering out all samples beyond the 95th percentile of the respective distributions as their variability was reported to be considerable^41^, especially for older studies. Moreover, while ADNI relied on predefined cutoffs, this procedure was not applied consistently across biomarkers. Finally, we computed an additional variable, the ratio between Aβ_1-42_ and Aβ_1-40_ and rescaled all features between 0 and 1 to convert data to a suitable scale for machine learning.

#### Neurological and Behavioral Assessments

We selected the total scores from all relevant neurological assessments. We subsequently applied min-max rescaling as described for CSF data preprocessing above. For the Neuropsychiatric Inventory Questionnaire (NPI-Q), we identified and marked all questionnaires that were left completely unanswered as missing. When a screening question was answered negatively, we assigned a severity score of zero to that answer. We then extracted only the 12 severity scores for each questionnaire, excluding the screening questions. We normalized the scores by dividing them by 3 (highest severity score) and constructed dense vectors as input to our models. An overview of all included data for each UC is shown in Table 2.

**Table 2.**
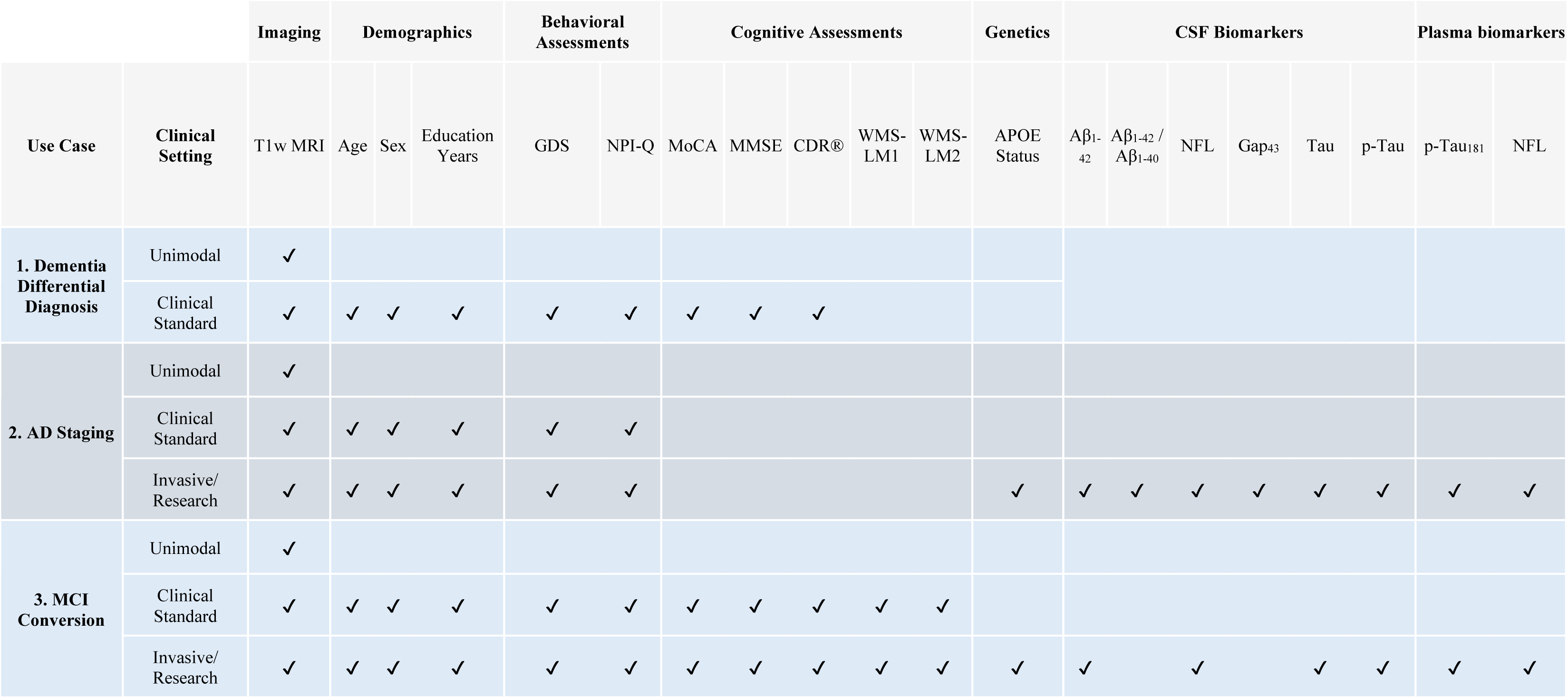
Overview of data included from ADNI, AIBL, and FTLDNI for each use case. . CSF = Cerebrospinal Fluid, WMS L1/L2 = Weschler Memory Scale (revised) Logical Memory 1/2, CDR = Clinical Dementia Rating, MMSE = Mini-Mental State Examination, MoCA = Montréal Cognitive Assessment, GDS = Geriatric Depression Scale, APOE = Apolipoprotein-E, Aβ = Amyloid-beta, GAP_43_= Growth-associated protein 43, p-Tau = Phospho-tau, NFL = Neurofilament Light

#### T1w MRI preprocessing

We included T1w MRI images from the ADNI 1-3-Go, AIBL, and NIFD databases in our analyzes by querying all relevant images (including both 1.5 and 3 Tesla acquisitions). We applied a deep neural network from ANTsPyNet to each T1w image for skull stripping. Each extracted brain was bias-corrected using ANTsPy and cropped to remove empty space. All images were then resampled, centered, and cropped to 128^3^ voxels so as to fit all images to a uniform size and isotropic voxel resolution of 1.7 mm^3^.

#### Cascaded Multimodal Mixing Transformers

To overcome data-missingness, we used PyTorch 2.0.1 to implement a Cascaded Multimodal Mixing Transformer (CMT), adapted from a previously described architecture^42^. Unlike other DL models that process all inputs simultaneously, CMT builds its final representation sequentially, enriching a learnable representation with information from different modalities step-by-step (Fig.1). In our implementation, we used this sequential nature to effectively exclude missing modalities from the processing chain instead of feeding zero-like embeddings to the blocks.

**Figure 1.**
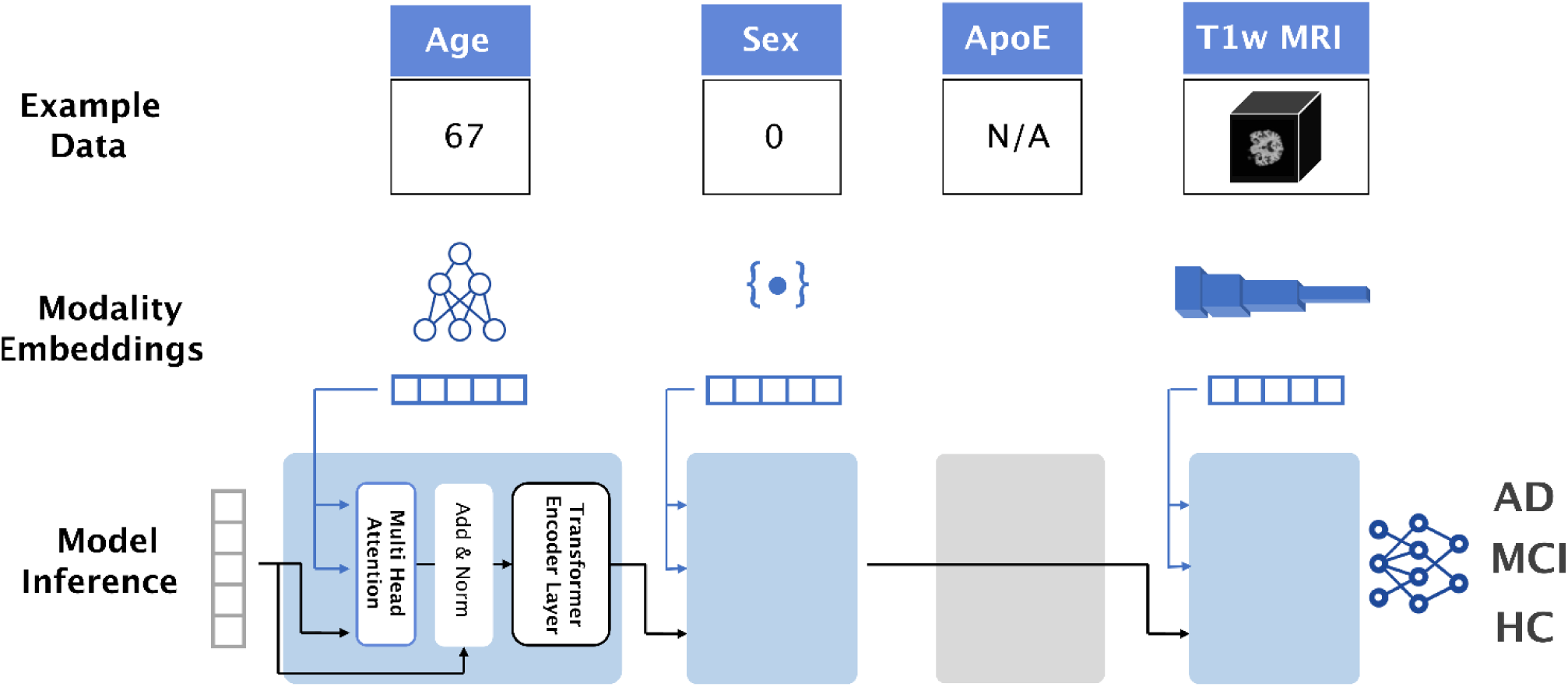
Cascaded Multimodal Transformer (CMT) Overview. Visual depiction of the inference approach adopted by the CMT. Each data point is individually embedded in a latent dimension. These embeddings then serve as keys and values in a cross-attention layer. Following normalization, a transformer encoder layer further processes the fused embedding. Each block follows the previous ones and integrates data only if the modality is available. The final embedding is then fed through a set of fully connected layers and mapped to probabilities to answer questions specific to each use case.

#### Modular Architecture

Our CMT consisted of blocks designed to convert data from different sources into a unified latent representation with a dimension of 256. Each block processed the output of the previous one by integrating additional information^42^. Each block contained three main components: an embedder, a cross-attention layer, and a transformer encoder. The embedders transformed our input modalities into modality embeddings with unique encodings that could be processed by the model. This allowed for the independent embedding of each modality separately. We encoded categorical data (i.e., APOE status and sex) with a lookup table, while ordinal data (such as assessment scores and age) were linearly projected to the latent dimension. Finally, we adopted a convolutional feature extractor as encoding architecture for volumetric MRI data. We applied cross-attention to merge each modality embedding with the latent one, sequentially. In this step, we translated the individual modality information (e.g. T1w MRI) to the transformer’s space using the modality embedding as the Key and Value and specified the Query as the model’s latent embedding to attend to the incoming data. Following each fusion, we applied self-attention and a forward network with a dimension of 1024, consistent with a standard transformer encoding layer^43^ (Fig. 1).

#### Modality Dropout

We leveraged the original modality dropout technique^42^ to account for bias originating from different data availabilities across modalities and to improve model robustness to missing data. Here, we randomly masked batches of data with *Not a Number* (NaN) according to a well-defined dropout rate (see further sections), thereby applying missingness upon existing data to make the model more robust to unavailable information, independently of the modality.

#### Data Sparsity and Bias

To account for possible biases common to models trained on complex, incomplete, and multimodal data, we took bias-correcting quality control steps. To account for biases resulting in discrepancies in data availability across studies (such as CSF biomarkers only being measured in ADNI), we used variables that were available for all the conditions selected for each use case (for example, we included CSF in UC2 but not in UC1). We also developed a new training framework referred to as Cascaded Training to deal with imbalanced classes with different amounts of missing data for each modality. Additionally, we used an established balanced accuracy metric to evaluate model decision-making better^44^.

#### Cascaded Training Framework

We developed the Cascaded Training Framework to address suboptimal learning outcomes caused by different data-missingness patterns across modalities and frequency of the diagnoses. The model’s blocks were sequentially and individually trained with a modality-specific loss weight computed on the class distribution within each modality. Once a block was trained, its weights were frozen, allowing for the next block to be added in sequence (Fig. 2). We conceptually separated the *training block* CMT_i_, which is the one being trained at a given iteration, from the *trailing blocks* CMT_j|j<i_ that appeared before CMT_i_ and provided context to the block being trained. In our experiments, for simplicity, we trained all categorical and ordinal blocks with a learning rate of 1E-5 for 20 epochs. To train the T1w MRI block, we raised the learning 5E-5 and used a linear learning rate decay schedule for 70 epochs. We used the Medical Open Network for Artificial Intelligence (MONAI) libraries^45^ to load NIFTI, rescale intensities between 0 and 1, apply random affine transformations and flip along the z-axis (transversal) to improve robustness and generalizability^46^. Data augmentation can result in data leakage between train and validation splits. Therefore, we used MONAI augmentation pipelines that work in a streamlined fashion to avoid this.

**Figure 2.**
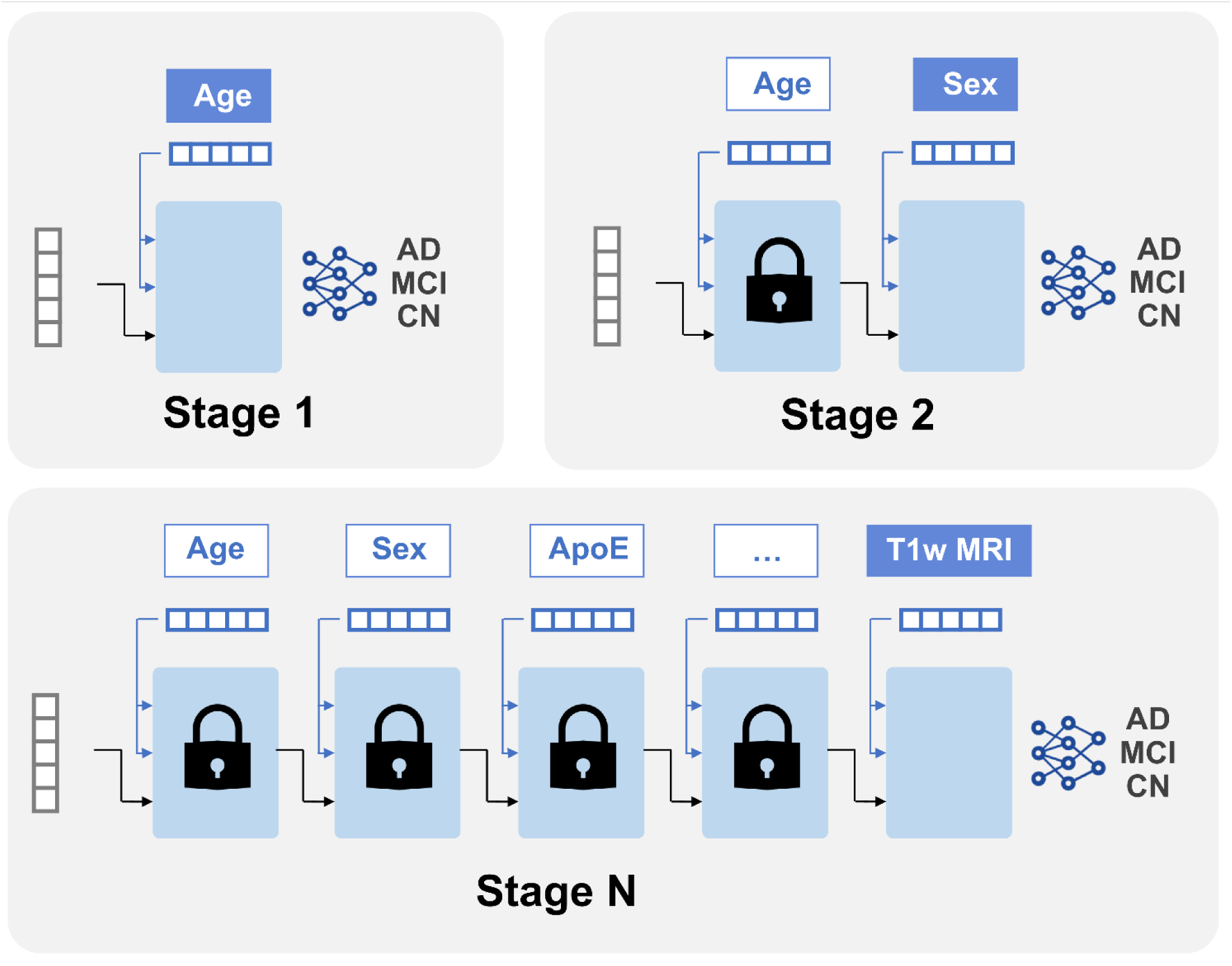
Cascaded training iteration. Representation of cascaded training showing trailing blocks that are “locked” as their weights are frozen. This example shows the integration of “Age” to the model at Stage 1, whose weight is subsequently frozen for the following integration of “Sex” (Stage 2) and ApoE (Stage 3). This process of integration and preceding freezing of weights is repeated with the inclusion of all further features to the model (Stage N). Despite the freezing of feature weights, they remain capable of processing the input and providing context to the training block, which is shown placed at the end of the chain.

#### Modality Dropout Computation

We used modality dropout on the *trailing blocks* to mitigate the bias associated with different data availabilities. Let’s denote with *CMT*_*i*_as the CMT’s Block that processes the modality *m*_*i*_. Before *CMT*_*i*_ began training, we filtered out from the original dataset all the observations where *m*_*i*_was missing, leading to a dataset 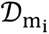 where the training modality was complete. We then computed the dropout to be applied on the trailing blocks in a way to have homogeneous amounts of missing data. Let’s denote with *r*_*target*_ the missing rate to be achieved in all trailing modalities. We removed from the target dropout the natural missing rate of the data (*r*_*data*_) in the identified subset. This strategy allowed us to avoid over-dropping of sparse modalities since their missing rate would have been close to the target dropout, hence resulting in a minimal additional dropout.

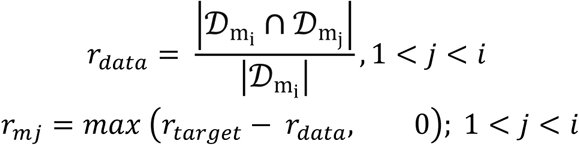

We adopted an expected dropout rate of 70% for tabular data, and subsequently raised it to 90% when training T1w MRI, in order to boost unimodal performance. Additionally, we implemented a *label-specific* modality dropout strategy to address potential biases that could arise from uneven data distributions across different classes, ensuring that no label was appearing more frequently than the others within the same trailing modality. This, in turn, prevents the model from associating data patterns too closely with specific classes, which can occur if certain modalities are disproportionately linked to specific classes (for example, in ADNI, CSF was acquired more often in MCI and CN). To achieve this, for each epoch, we determined the extent of missing data for each modality within each label. For each modality, we identified the highest rate of missing data across all labels. Then, for labels with lower rates of missing data for a given modality, we artificially increased the amount of missing data to match the highest rate observed. We accomplished this by randomly selecting a subset of data points to be masked as if they were missing. In the hypothetical situation where a modality was present exclusively in one diagnosis, this dropout would have assumed the value of 1 (total dropout) for the other classes, resulting in no learning regarding that modality.

#### Explainability

We relied on graphical and quantitative methods to establish the origin of our model’s errors and to characterize the response of our model to different modalities. We adopted t-distributed stochastic neighbor embedding (t-SNE)^47^ to visualize high-dimensional features learned by the embedders of each block in a 2-dimensional space. We also observed that the input-output relationship of cross-attention carried value in explaining how the model was responding to different modalities, independently of their nature. We propose then a new cross-attention-based explainability metric, Cross-Modal Fusion Norm (CMFN), as follows:

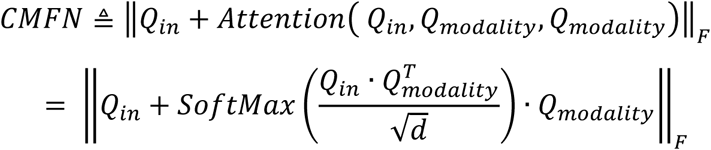

In this formulation, Q_in_ is the multimodal embedding, Q_modality_ is the embedding about to be merged and *d* is the latent dimension. We hypothesized that Q_in_ encapsulates information about the patient, which is progressively enriched through the information flow, while Q_modality_ provides contextual information about the input modality. As a result, the proposed metric aims to describe the modality-specific cross-attention response given the participant’s characteristics which are comprehensively stored in *Q*_*in*_. We computed the average CMFN for each class to understand important mechanisms across features, and we plotted how this metric was reacting at different values within the same modality to get insights on what was considered meaningful. For high-dimensional modalities, we relied on t-SNE to reduce the dimensionality of the embeddings and highlighted the coordinates by CMFN magnitude as shown in Fig. 5.

#### Post-Hoc Analysis

Following model training on different folds, we evaluated each on its respective validation fold. This cross-validation process provided performance metrics and confidence intervals, which we aggregated to evaluate the model’s overall performance across the dataset, enabling global confusion matrix creation, post hoc analysis, and attention analysis. We also analyzed misclassified data to uncover sources of error. Where we could not plot the input values to interpret where the model misclassified, we again adopted t-SNE as a dimensionality reduction tool and colored correct and incorrect predictions with different markers.

## Results

Comparison of our model performance to state-of-the-art results obtained from other literature (see supplementary section 3) show varying results for each UC (Fig. 3), with poorer performance in the unimodal approach of UC1 and equal or improved performance in UC2 and UC3, particularly in the Clinical Standard and Invasive/Research multimodal modeling approaches. For UC1, we report two performances: for the full differential diagnosis, 5-classes prediction (classes being: CN, AD nfvPPA, bvFTD, and svPPA; chance level 20%) and when comparing with the literature, for a 3-classes prediction (CN, AD, FTLD; chance level 33%) obtained by grouping the FTLD subclasses predictions to one new class. This aggregation was necessary to provide a robust comparison with existing work.

**Figure 3.**
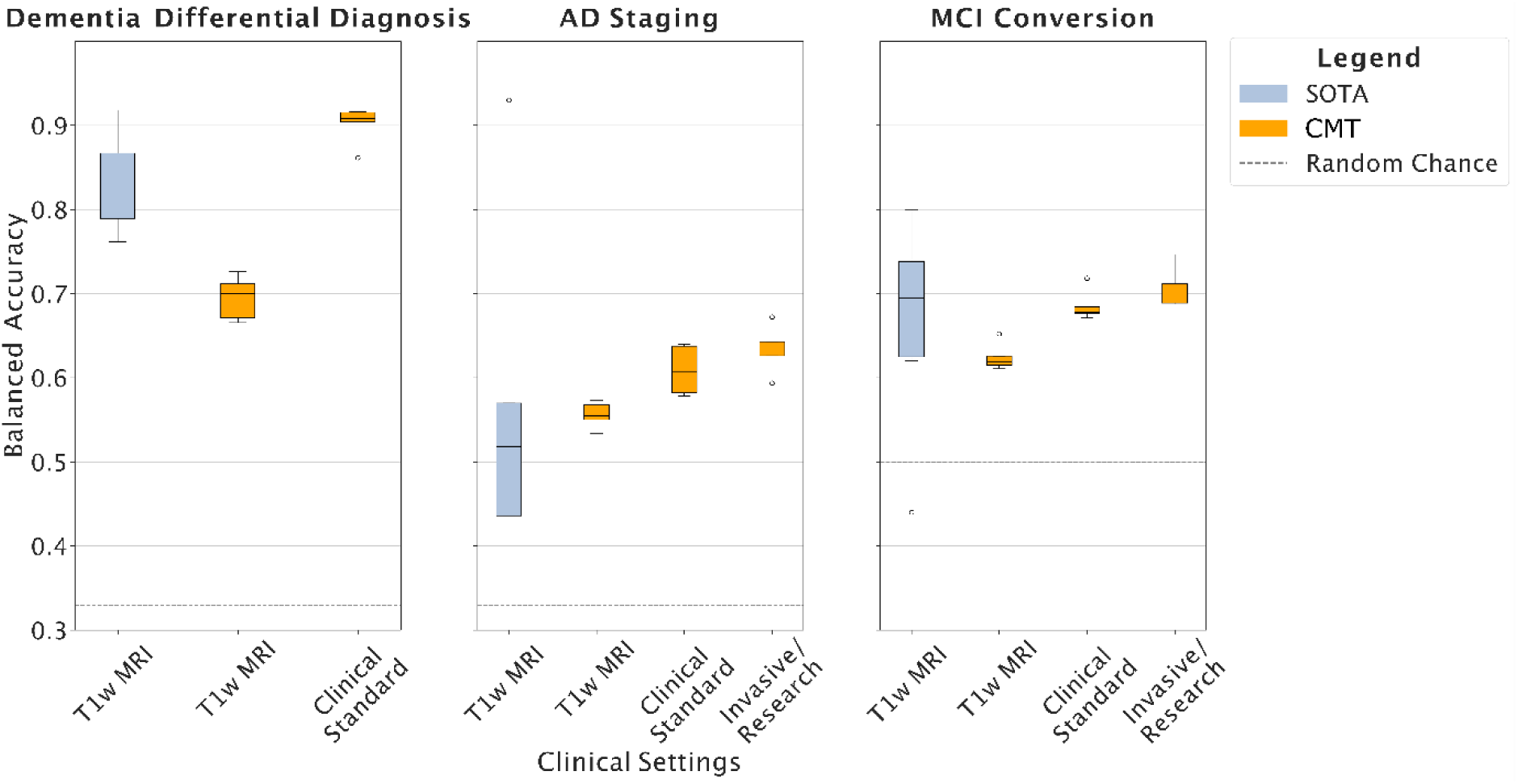
Performances obtained in different Use Case Scenarios and Clinical Settings. Results show that multimodality (i.e. adding more forms of data to the model) enhanced model accuracy in each Use Case. For UC1, aggregated performances are shown (CN vs AD vs FTLD). In this UC, unimodal performance yielded a lower balanced accuracy compared to previous literature, while multimodal modeling increased balanced accuracy. Results from each of UC2 and UC3 show gradual increases in balanced accuracy with increasing modality, improving on results obtained from previous studies. SOTA = State-of-the-art, CMT = Cascaded Multimodal Mixed Transformer.

### Use Case 1: Differential Dementia Diagnosis

#### Unimodal

Using only T1-w MRI, our model achieved a balanced accuracy of 61.1±4.2% (confusion matrix in Fig. 4A; chance level 20%). As previously described, predictions for FTLD subclasses were aggregated into one class to facilitate comparison with the literature. This resulted in 69.5± 2.6%. The Receiver Operating Characteristic (ROC) showed a robust area under the curve (AUC) for all classes, with the minimum AUC being 0.79 for AD versus all the other classes. However, this was accompanied by large confidence intervals (ROC curves color bands in Figure 4A). A t-SNE reduction of the generated features allowed the identification of clear clusters for svPPA and bvFTD. Still, nfvPPA was not well defined in the geometric space with major overlaps between bvFTD and CN. A partial overlap was also observed between AD and CN features.

**Figure 4.**
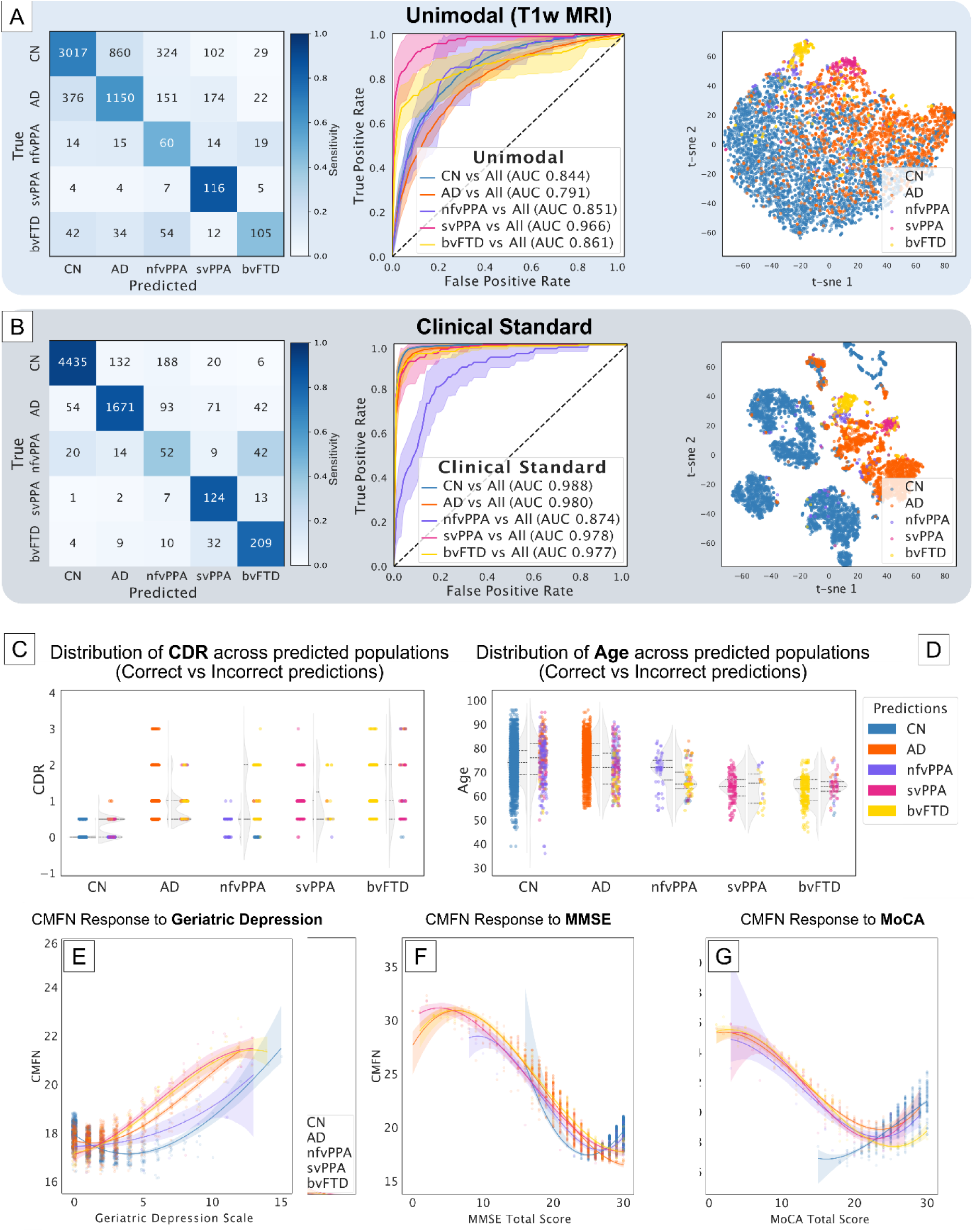
Results of Diagnosis Classification (UC1). Confusion Matrix, ROC Curves (Mean FPR, and TPR ±1SD computed across 5 validation folds), and t-SNE reduction of the final embeddings for the unimodal setting (A), and the Clinical Standard setting (B). D-E) Error Analysis showing how MMSE (D) and Age (E) are distributed among misclassified and correctly classified individuals for each condition. F-H) Cross-Modal Fusion Norm (CMFN) magnitude at each value of MoCA (F) MMSE (G) and Geriatric Depression Scale (H) colored by label. SD = Standard Deviation, CN = cognitively normal, AD = Alzheimer’s disease, nfvPPA = non-fluent variant primary progressive aphasia, svPPA = semantic variant primary progressive aphasia, bvFTD = behavioral variant frontotemporal dementia, MMSE = mini-mental state exam, MoCA = Montréal cognitive assessment, CDR = clinical dementia rating.

#### Clinical Standard

When adding cognitive, behavioral, and demographic Clinical Standard information, the model achieved a balanced accuracy of 75±3.8% (confusion matrix in Figure 4B). Aggregation of predictions resulted in 90.1±2.0% balanced accuracy, marking a significant increase in disease classification accuracy compared to the unimodal approach. Sensitivity towards nfvPPA decreased due to substantial difficulty in distinguishing it from bvFTD. ROC analysis for the multimodal approach showed a marked improvement in AUC across all predicted classes, indicating better overall performance (Figure 4B). Note that nfvPPA retained a high AUC but in comparison with other syndromes, yielded the lowest AUC improvement. Additionally, the variability of performance across folds was qualitatively reduced. A t-SNE dimensionality reduction of the multimodal embeddings showed a complex space organization resulting from different modality values. The space showed clusters characterized by inferior variability compared to the unimodal scenario, particularly with AD being significantly better separated from CN.

#### Error Analysis

We analyzed model misclassifications and discovered the model generated false negatives when cognitive performance was better in AD, nfvPPA, and bvFTD. Likewise, the opposite was also true (Fig. 4C). When age was considered, we found most of the AD misclassifications happening in subjects above 70 years old while most of the errors below this threshold were happening with FTLD (Fig. 4D). In terms of psychological assessments, we observed abnormal depression levels in cognitively normal volunteers, thereby resulting in misclassification as AD, nfvPPA, and bvFTD.

#### Explainability

The Cross-Modal Fusion Norm (CMFN) analysis provided insights into how different modalities contributed to the classification (Fig. 4E-G, 5A). For cognitive assessment scores such as MoCA and MMSE, the model placed higher attention on lower scores. Higher scores were also attended to in cognitively normal volunteers (Figure 4F, 4G). Geriatric Depression Scale (GDS) scores also showed complex responses in that cognitively normal people received more attention for low scores but, as the score increased, other dementia classes overtook the attention over CN (Fig. 4D) showing that the metric can capture a stratified response exhibited by the model. The NPI-Q and T1-w MRI data showed similar patterns, with higher attention given to svPPA and bvFTD classes (Fig. 5A). Furthermore, when CMFN was inspected in MRI features, we found that elevated attention was placed on features belonging to FTLD conditions while AD features received much less attention in comparison (Fig. 5A).

**Figure 5.**
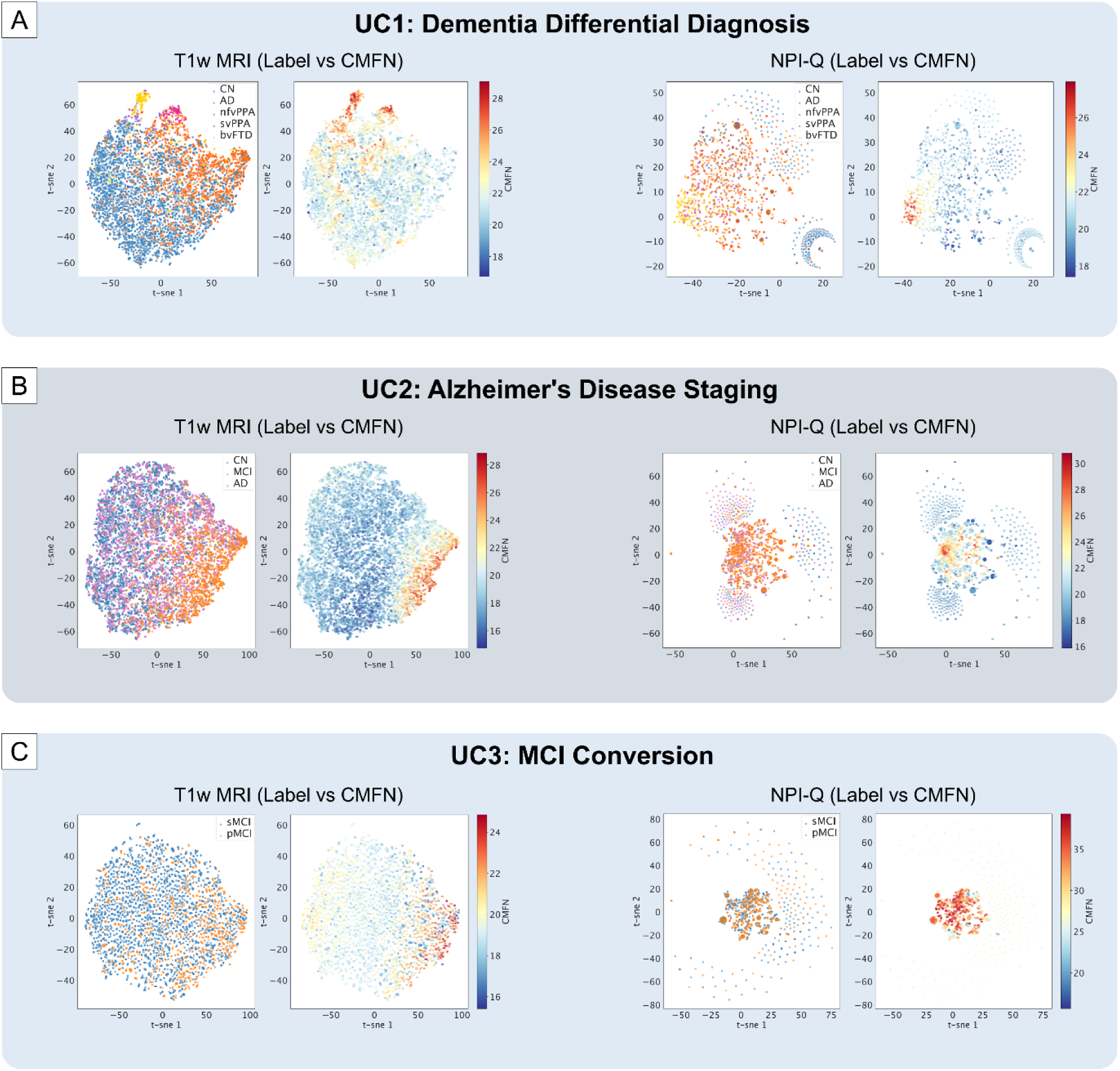
High Dimensional Attention Analysis on T1w MRI and NPI Features. Each sub-figure shown in pairs: on the left the features are colored by condition (or label), and on the right the CMFN is highlighted. This frame enables the comparison of which condition received the most attention. (A) Analysis results for the Differential Diagnosis task, (B) for AD Staging, and (C) for MCI conversion where, however, the CMFN was affected by convergence issues in the early layers. NPI-Q = Neuropsychiatric Inventory Questionnaire, CMNF = Cross-Modal Fusion Norm, t-SNE = t-distributed Stochastic Neighbor Embedding.

### Use Case 2: AD-Staging

#### Unimodal Approach

When the model input was limited to T1w MRI data, the achieved balanced accuracy was 55.6±1.4% (chance level 33.3%), as reflected in the ROC curves (Fig. 6A), with specific challenges in detecting MCIs reliably. This was especially evident when looking at the t-SNE plots showing MCI overlapping with the other two classes (Fig. 6A). Note that MCI was *not* stratified into converters and non-converters to AD when training the model for this use case.

**Figure 6.**
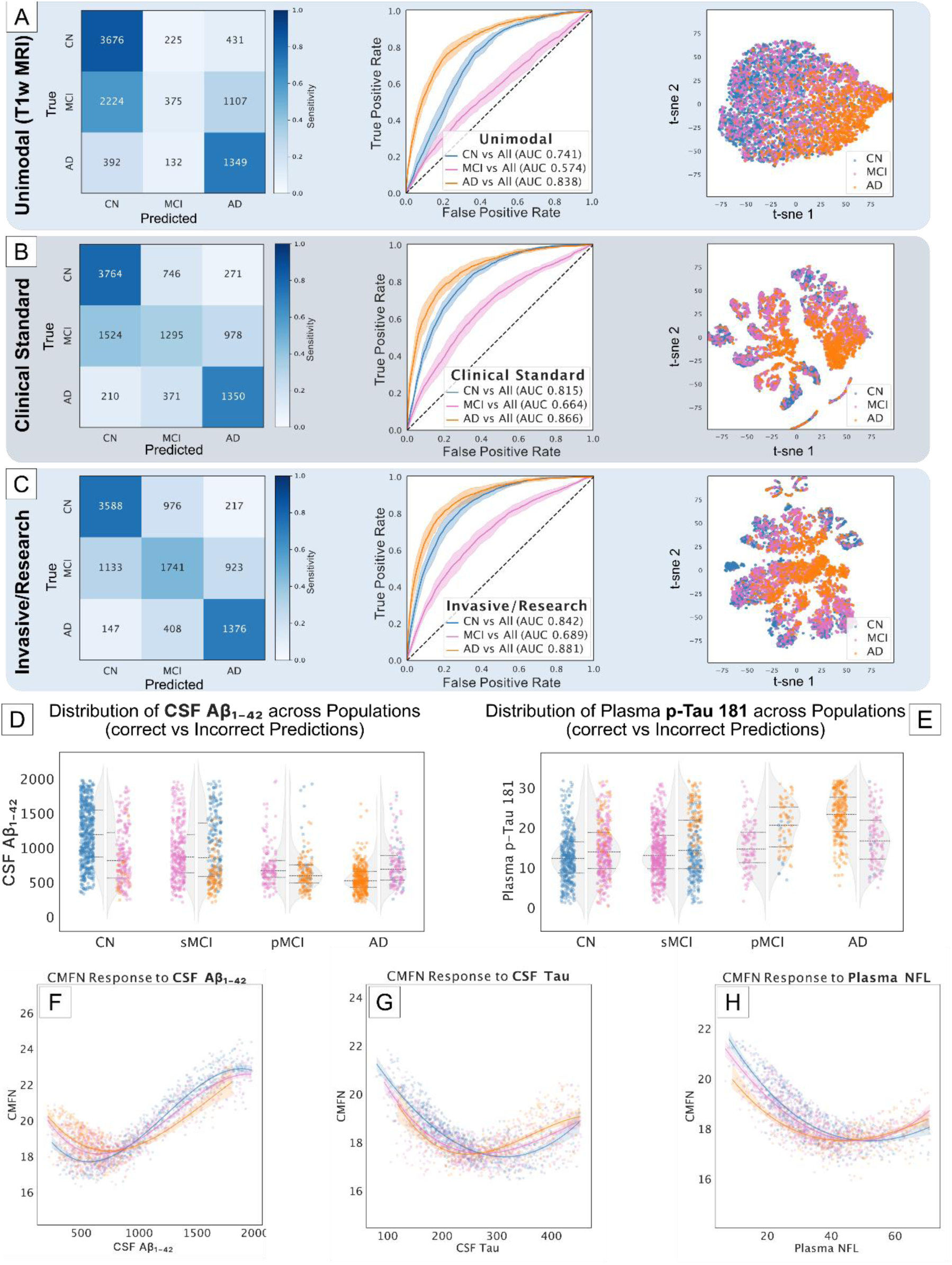
Results of AD Staging Modeling (UC2). Confusion Matrix, ROC Curves (Mean FPR and TPR ± 1SD computed across 5 validation folds), and t-SNE reduction of the final embeddings for (A) the unimodal, (B) Clinical Standard, and (C) Invasive/Research settings. Results show increased balanced accuracy (from 55.6% to 64.1%) and an increase in MCI sensitivity (from 10% to 56%). Error analysis shows Aβ_1-42_ (D) and plasma p-Tau (E) distributions. (F-H) CMFN magnitude for each value of Aβ_1-42_ (F) Tau (G) and Plasma NFL (H) by diagnosis. SD = Standard Deviation, CN = cognitively normal, sMCI = stable mild cognitive impairment, pMCI = progressive mild cognitive impairment, AD = Alzheimer’s disease, CSF = cerebrospinal fluid.

#### Clinical Standard

With the further inclusion of behavioral assessments and demographic data, the balanced accuracy increased to 60.9±2.6% (Fig. 6B). MCI sensitivity showed the biggest change since it improved from 10% to 40%. ROC analysis for this multimodal approach showed an improvement in AUC across all predicted stages, indicating a better overall performance (Fig. 6B).

#### Invasive/Research

In the most comprehensive setup including all multimodal data available, the balanced accuracy reached 64.1±2.9%. This setup provided the highest accuracy and sensitivity (Fig 6C). ROC analysis further supported this finding, with the highest AUC values observed in this setup, indicating superior classification performance. It is important to frame the improvement within the context of the sparsity of CSF data, which averaged 80.6% missingness in our dataset. Interestingly, we found that the sensitivity to AD and CN remained nearly unchanged across unimodal and multimodal settings since, also in this last case, the primary driver in accuracy was found in MCI sensitivity which improved by 6% compared to the Clinical Standard (as can be inferred from the confusion matrices in Fig. 6A-C).

#### Error Analysis

Though we trained our model in a three-class setting, we analyzed the differences in misclassifications in the pMCI and stable MCI (sMCI) populations separately to understand better when the model was failing (note that pMCI and sMCI correspond to converters and non-converters from MCI to AD in the literature^48^). We found differences in misclassification between the two groups: pMCI were overall more readily detected as MCI, with a sensitivity of 56%, however, 75% of misclassifications happened towards AD. On the other hand, sMCI proved to be harder to recognize, with a sensitivity of 47% and a disproportion in misclassifications (66%) towards the normal class. Misclassified cognitively normal participants were generally older and less educated in the case of MCI and AD predictions. Behaviorally, misclassified CN individuals had more severe GDS scores when misclassified as MCI and AD. Important differences across distinct prediction groups were especially found in biomarkers where observations predicted as AD had higher Tau, p-Tau, Plasma p-Tau 181, Plasma NFL and lower Aβ_1-42_, and CSF Aβ_1-42_/Aβ_1-40_; analogous but opposite patterns were observed for mistakes towards CN (lower Tau, p-Tau, higher Aβ_1-42_ and CSF Aβ_1-42_/Aβ_1-40_) (Figures 6D, 6E). In all biomarkers, pMCI distributions were more similar to AD than the stable group. This behavior was also mirrored by the sMCI group, where mistake clusters for the CN and the AD groups showed opposite patterns in CSF and plasma biomarkers (Figures 6D, 6E).

#### Explainability

The CMFN indicated plasma and CSF biomarkers as the most impactful information, with Aβ_1-42_ being the most important. The CMFN analysis revealed distinct patterns for various biomarkers. For Aβ_1-42_, the CMFN was higher for AD and MCI at lower levels and higher for CN at higher levels (Figure 6F). CSF Tau, p-Tau, NFL, and plasma NFL showed higher attention for cognitively normal individuals at lower biomarker levels compared to AD and MCI, with the trend reversed at higher values (Figures 6G, 6H). The model also showed significant attention to the APOE genotypes, particularly (2,3) and (4,4). High-dimensional attention analysis through t-SNE revealed that MRIs belonging to AD featured high CMFN, while the metric dropped in the middle region where the overlap between AD and CN was higher (Figure 5C).

### Use Case 3: MCI Conversion

#### Unimodal Approach

Using only T1-w MRI for prognosis yielded moderate results regarding balanced accuracy (63±1.5%; chance level 50%) (Fig. 7A) and AUC (0.673) which were reflected in the t-SNE feature space where no clear separation emerged.

**Figure 7.**
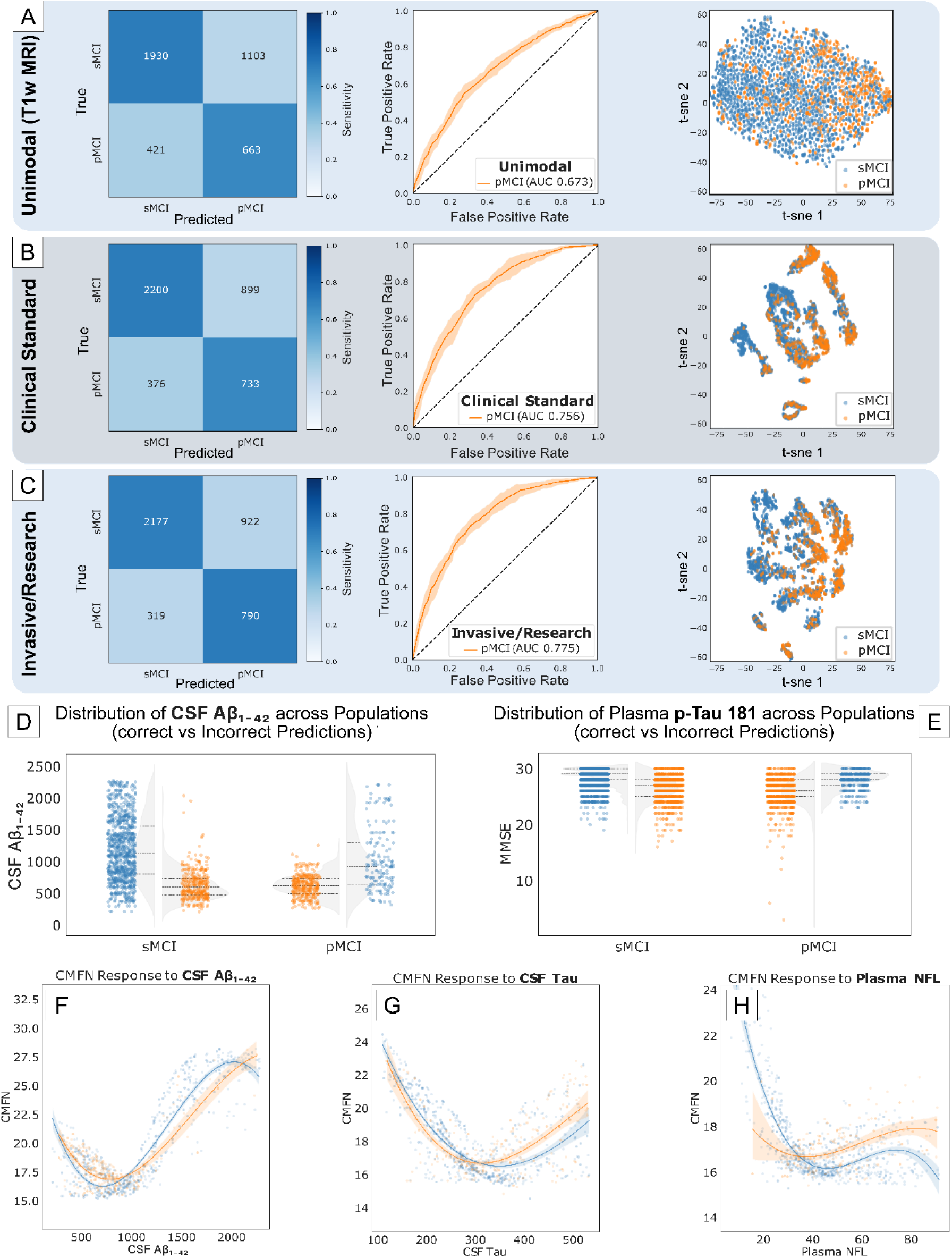
Prediction of MCI conversion (UC3). (A) Confusion Matrix of performance and (B) comparison of generated embeddings across each setting. (C) ROC Curves, Mean FPR, and TPR ±1SD computed across 5 validation folds. (D-E) Error Analysis showing Aβ_1-42_ (D) and MMSE (E) distributions in misclassified and correctly classified individuals. (F-H) CMFN magnitude at each value of Aβ_1-42_ (F) p-Tau (G) and plasma NFL (H) colored by label. SD = Standard Deviation, AUC = Area under the curve, s/pMCI = stable/progressive mild cognitive impairment, CSF = cerebrospinal fluid, NFL = neurofilament light.

#### Clinical Standard

Incorporating cognitive and behavioral scores alongside demographics notably boosted the model performance, yielding a 69±1.8% balanced accuracy (Fig. 7B). We also observed this improvement in the ROC Curve, which showed an improved AUC of 0.756.

#### Invasive/Research

In line with other UCs, the accuracy and AUC peaked in the most comprehensive setting (Fig. 7C). Incorporating CSF, plasma, and APOE biomarkers allowed for improved confidence in the predictions which was reflected in the AUC (0.774) and balanced accuracy (70.8±2%).

#### Error Analysis

Analyzing where the model struggled in predicting the conversion to future AD highlighted many possible reasons for misclassification. Regarding demographics, we observed that persons misclassified as pMCI had significantly higher age, and lower education and that cognition played a key role given that misclassified pMCI had cognitive scores towards the lower end of the distribution. CSF and plasma biomarkers revealed similar results since the biomarker profile of the misclassified subgroup overlapped with the one from the antagonist group (Fig. 7D, 7E). Finally, in all settings, we discovered a longer time to conversion between the observations of false negative pMCI, as shown in Figure 8.

**Figure 8.**
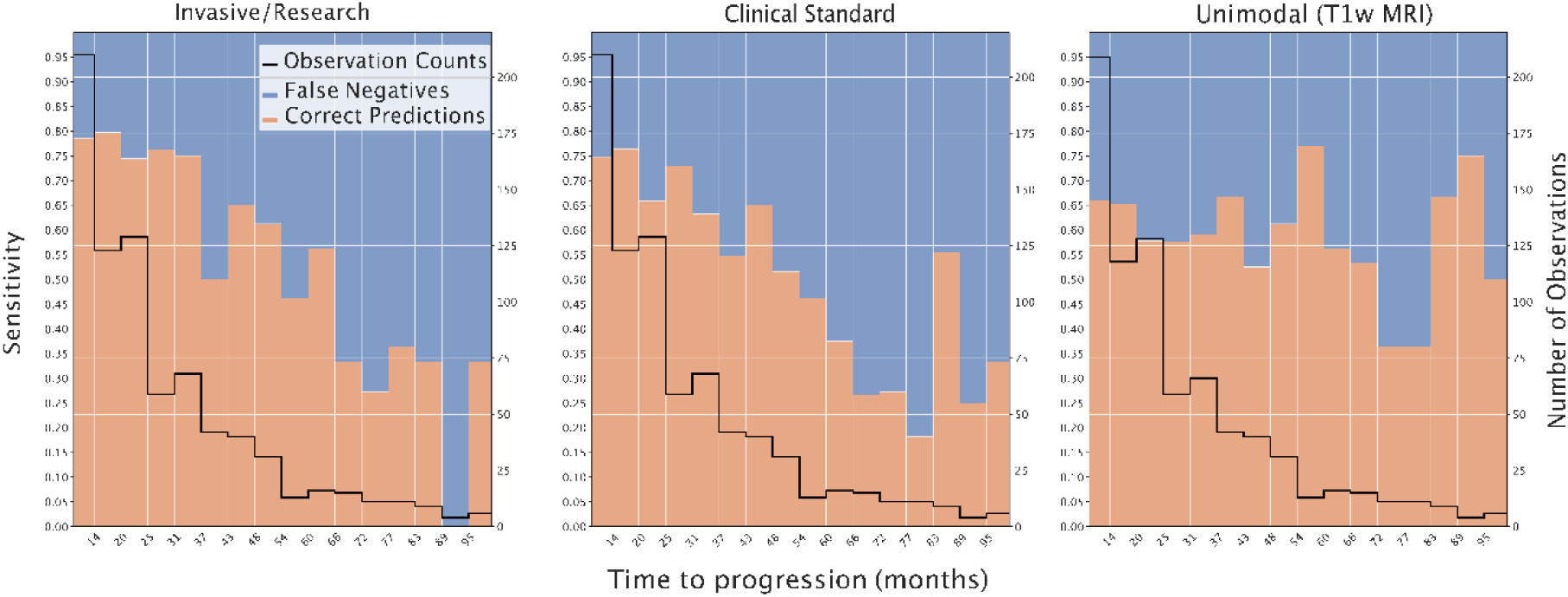
Sensitivity to AD progression in the prognosis of MCI conversion. Multimodal models exhibit higher sensitivity for shorter timespans, which also show a decrease over time. The unimodal modeling approach shows a less steep decline though results show overall worse specificity. AD = Alzheimer’s disease.

#### Explainability

We observed an anomalous behavior in the CMFN for demographic modalities where the model did not effectively make use of this information, causing local divergence in the early blocks. This resulted in an inflated CMFN that made the interpretation of important features impossible. The inflated CMFN issue primarily affected the modalities placed at the beginning of the CMT chain but gradually dissipated in later blocks. In the latter blocks, however, we observed that the CMFN was elevated in sMCI compared to pMCI. We observed the opposite pattern, however, for Aβ_1-42_ whereby lower levels were more attended in sMCI (Fig. 7 F-H).

## Discussion

Both preclinical and clinical data are complex in nature and are often siloed. These data are thus difficult to integrate. To assess the validity of Deep Learning (DL) technologies in addressing this in the context of dementia research and healthcare, we have applied a CMT architecture to three neurodegenerative diseases Use Cases (UCs) using data from the six open-source data sets. In UC1, we evaluated TelDem first using a unimodal and then a multimodal modeling approach, in the differential diagnoses of cognitively normal adults, AD, and three subclasses of FTLD. In UC2, we evaluated the model in the prognostic staging of AD, comparing model performance firstly in an unimodal approach, secondly in a clinical standard approach, and finally in a multimodal approach. In UC3, we investigate disease progression, testing the model’s ability to distinguish between progressive and stable MCI. Overall, our results show that the addition of multiple modalities improves model accuracy compared to the modeling of unimodal data.

### Use Case 1 – Dementia Differential Diagnosis

Compared to other unimodal automated solutions^49,50^, TelDem achieved superior balanced accuracy (0.901) (CN vs AD vs FTLD) despite underperforming when only using MRI compared to previous studies^51–53^. The incorporation of multiple data modalities enhanced diagnostic accuracy by 14% and improved confidence of the predictions, with notable increases in the AUC across most diagnostic groups. This improvement highlights the system’s ability to leverage diverse diagnostic data effectively, though it exhibited a multimodal trade-off, particularly in nfvPPA, where a more frequent confusion with the behavioral trait suggested an over-reliance on behavioral assessments for this diagnosis. While sensitivity decreased, the AUC remained the same however, which indicates that the model is still capable of ranking positive cases with the same decision threshold. Additionally, not all participants underwent MRI at all encounters, which could moderate sensitivity in situations where MRI is critical, such as nfvPPA.

There are several hypotheses regarding why the model struggled with the nfvPPA diagnosis. First, despite broad clinical and anatomical differences, both bvFTD and nfvPPA show similar focal neurodegeneration in the insulae^54^, longitudinal atrophy in dorsolateral and prefrontal regions^55^, and elements of tau histopathology^56^. Secondly, nfvPPA shows locally unspecific and limited brain atrophy in comparison to svPPA, making detection with MRI challenging^57^. Including assessment of language comprehension could possibly improve nfvPPA classification. Additionally, nfvPPA had the least available data and, while our model can handle this, it is possible that a limited training size in conjunction with these other ambiguating factors played a role in model underperformance.

In the context of explainability, CMFN analyzes revealed that T1-w MRI and the NPI-Q, which is specifically designed to assess psychopathology in dementia^58^, were the most critical in model decision-making for UC1. One explanation for this relates to the variety of behavioral and psychological symptoms that occur in dementia (BPSD)^59,60^ and how these characterize different dementia disorders^61^. For example, changes in eating habits are reported to be more frequent in FTLD than in AD^62^ while delusions occur more in AD^63^. However, it’s also important to highlight that BPSD symptomatology is not static^64^ and there may be overlap across FTLD subtypes. This dynamic nature of BPSD could have contributed the conflation of nfvPPA with bvFTD.

Secondly, MRI was critical in differentiating FTLD subtypes, which is unsurprising given that MRI allows for the identification of AD-specific patterns of atrophy or of different subtypes of FTLD^65,66^. However, the model may have also relied on demographics and cognitive scores to classify AD as less attention was attributed to MRI for this diagnosis compared to FTLD. Interestingly, CMFN also showed that the CDR received less attention at intermediate values, which suggests that the model relied on this assessment to rule out dementia, rather than to distinguish between different subtypes. This is consistent with the literature, given evidence that the CDR has a limited utility in discriminating FTLD from AD^67^. For this reason, efforts to integrate more extensive neurological domains to the CDR are underway^68^. Moreover, model responses to the same information varied depending on the diagnosis. For example, CN participants and persons with dementia showed different response patterns in cognitive scores, suggesting the model can downweigh outlying information, such as low cognitive scores. For GDS ratings, we observed the opposite where, for a total score of zero, CN participants received the most attention. Attention at higher scores was dominated by svPPA, bvFTD, and AD.

### Use Case 2 – AD Staging

Regarding UC2, our system underperformed in unimodal scenarios when solely using MRI to stage AD. While this could be attributable to the limited image resolution, multimodal modeling including demographics and behavioral scores boosted performance, reinforcing the importance of including broader clinical metrics in the diagnostic process. Including these biomarkers also improved both AUC and sensitivity. However, errors with this diagnosis were still a major source of inaccuracy. This outcome was nevertheless anticipated due to the heterogeneous nature of MCI, which does not always manifest clearly in biomarker profiles^69^, possibly leading to the observed misclassification of pMCI as AD. While the inclusion of CSF and plasma biomarkers did show improvements in model performance, these gains were limited by the scarcity of these biomarkers in our dataset. While this would suggest that addition of these more fine-grained measures could help with classification, it is possible that increased availability could yield only minor improvements, given that diagnostic criteria for AD often rely heavily on cognitive readouts only^70^. Hence, while biomarkers provide valuable information that help with predicting cognitive decline, their potential to fully resolve misclassification remains constrained by both availability and the limitations of the current AD diagnosis^71^.

Similar to UC1, we also aimed to explore model explainability. Our CMFN analysis revealed that CSF data received, on average, the most attention with Aβ_1-42_ being the most discriminative (along with its ratio). Conversely, Aβ_1-40_, tau, NFL, and gap_43_ were received less attention. This aligns with evidence showing that (i) Aβ_1-42_ is more diagnostically relevant compared to Aβ_1-_ ^72^, that changes in CSF Aβ occur early in the course of preclinical AD^73,74^, and that Aβ is a reliable CSF-based diagnostic marker of AD^75,76^. MRI also received high attention with regards to model decision making but primarily in AD, suggesting that the model was relying on other information to exclude pathology. CMFN analyzes also showed lower explainability with larger overlap between AD and CN. This may be due to MCI being an intermediate stage between normal cognition and AD^77^. Similar to cognitive scores in UC1, the model’s attention responded variably to CSF biomarker levels, suggesting an ability to contextualize biomarker data within the broader diagnostic picture. The model also focused more on APOE genotypes (e.g., 2,3 and 4,4), which again is consistent with literature showing the role of APOE genotype, in particular, in influencing AD risk^78,79^.

### Use Case 3 – MCI Conversion

Predicting the progression of MCI to AD (UC3), our results suggest an enhanced prediction accuracy when using multimodal DL to integrativley model all available diagnostic data. Although the unimodal approach underperformed relative to benchmarks cited in other literature, our multimodal model returned an accuracy of over 60%. The integration of cognitive scores was particularly effective, boosting prediction confidence and overall accuracy from 61% to 69%. The addition of CSF markers and APOE genotyping further increased the sensitivity for detecting progression to AD, achieving a global balanced accuracy of 71%. This observation would again buttress the diagnostic role of CSF biomarkers and could explain the small improvement from the clinical standard model to the full multimodal model as being a result of large amounts of unavailable CSF data.

We also observed higher sensitivity for faster conversion using all multimodal data compared to unimodal MRI. In fact, our model proved able to sustain above 75% sensitivity and up to 80% during the 35 months before receiving an AD diagnosis and stabilizing at around 30% six years before conversion. However, in the clinical standard scenario, sensitivity dropped faster suggesting that biomarkers indeed drove the gain. Interestingly, we did not observe the same drop in sensitivity when relying on unimodal MRI. We interpret this finding with caution, however, as it is likely due to an increased false-positive rate. Specifically, while the model demonstrated better sensitivity over longer periods, it did not perform as well over shorter periods, indicating that the increased sensitivity might be accompanied by a higher rate of incorrect predictions. In other words, the model identified stable individuals as likely to progress to AD when they were not, thus inflating the sensitivity metric at the cost of specificity. This is a known issue when relying solely on MRI data, for example, as many individuals with MCI exhibit brain atrophy suggestive of AD progression without progression within the expected timeframe^80^. This discrepancy could explain why the model did not achieve exceptionally high sensitivity at shorter prediction intervals.

#### Misclassification

Additionally, we also aimed to understand cases in which our model did not accurately classify participants. For example, in UC3 we observed that some sMCI observations were erroneously predicted as pMCI. This observation would seem to challenge the accuracy of our model. However these participants exhibited CSF Aβ_1-42_ levels and other biomarker profiles more closely resembling those of pMCI participants. This is notable given that diagnoses in the ADNI sample were made without reference to biofluid markers^81^. Previous research suggests that use of only cognitive scores can generate inconsistent results, with low memory scores, for example, being common in older individuals though varying significantly across different populations^82,83^. Our CMFN analysis shows that our model attributed high importance to CSF markers in its decision making, suggesting that these misclassifications may represent a more fine-grained labeling than that provided by the ADNI data set, rather than poor model performance per se.

#### Limitations

These interpretations, however, should be considered in the light of several limitations. First, the absence of multimodal neuroimaging in our approach could limit the power of our modeling, particularly given recent landmark studies that have made impressive strides forward in integrating and modeling multimodal imaging data^84^. Moreover, both Aβ and tau PET could be particularly informative given the associations of these metrics with cognitive decline and the robust predictive ability of PET-assessed tau accumulation for disease progression^85–88^. We chose only T1-w, however, first for feasibility, and second to better match the most widely available clinical routines. Second, we applied only cross-sectional predictions. Modeling of biomarkers longitudinally and capturing their multimodal interactions would likely enhance model performance, particularly in UC2 and UC3. We focused on cross-sectional data, however, as longitudinal modeling would unavoidably expand model complexity, thus obscuring interpretability and explainability. Third, CMFN analysis is possibly misleading when models are trained on less informative data, such as with the use of demographics in the early training stages of UC3. This likely resulted in a lack of divergence of initial blocks, inflating the CMFN norm. Hence, caution is needed when associating this metric with “feature importance”. Placing such variables later in the data integration sequence could reveal clearer patterns, suggesting that the order of data integration obscures model interpretability.

Fourth, we observed variability in reported metrics compared to other studies which could be due to data leakage. Leakage can bias evaluation of real-world model performance and recent literature suggests data leakage may explain why DL models achieve exceptional performance on one dataset but fail to replicate on another^89^. However, it must be noted that we deliberately specified our unimodal baselines to include features that showed less leakage through more robust methods^90^ or provided results based on independent datasets. Fifth, we did not account for medication use, which has been shown to modulate cognitive responses in ADNI and other similar data^91^. Future studies aiming to replicate our results should include this information to assess the degree to which medication use may affect model performance. Finally, we did not account for patient race or ethnicity. This is of critical importance in ensuring inclusion of people that are typically underrepresented in clinical research^92^. To ensure that TelDem is applicable to *all* patients, future studies should avail of ongoing efforts^93^ to include patients spanning a spectrum of racial, linguistic, and geographic backgrounds.

## Conclusion

DL applications represent an unprecedented opportunity to accelerate dementia research patient care. Nevertheless, stringent validation of DL-based systems is required. Here, we have deployed, evaluated, and assessed a multimodal DL architecture in the context of three UCs. Our results show that multimodality (i.e. the addition of diverse modalities) significantly improves disease classification, staging, and progression from MCI to AD, over unimodal modeling (the sole use of T1-w imaging). Moreover, model explainability revealed that CSF markers of Aβ contributed heavily to model decision-making, thus further supporting model validity. While additional research including Aβ and tau PET and more diverse patient data are needed, our results take a much-needed step in showing the advantages inherent to implementing DL research and clinical care. In conclusion, our results represent a new horizon for the efficient implementation of personalized treatments in dementia, thus providing a promising platform for enhancing research and patient care.

*Data used in preparation of this article were obtained from the Alzheimer’s Disease Neuroimaging Initiative (ADNI) database (adni.loni.usc.edu). As such, the investigators within the ADNI contributed to the design and implementation of ADNI and/or provided data but did not participate in analysis or writing of this report. A complete listing of ADNI investigators can be found at: http://adni.loni.usc.edu/wp-content/uploads/how_to_apply/ADNI_Acknowledgement_List.pdf

** Data used in the preparation of this article was obtained from the Australian Imaging Biomarkers and Lifestyle flagship study of ageing (AIBL) funded by the Commonwealth Scientific and Industrial Research Organisation (CSIRO) which was made available at the ADNI database (www.loni.usc.edu/ADNI). The AIBL researchers contributed data but did not participate in analysis or writing of this report. AIBL researchers are listed at www.aibl.csiro.au

*** Data used in preparation of this article were obtained from the Frontotemporal Lobar Degeneration Neuroimaging Initiative (FTLDNI) database (4rtni-ftldni.ini.usc.edu). The investigators at NIFD/FTLDNI contributed to the design and implementation of FTLDNI and/or provided data but did not participate in analysis or writing of this report (unless otherwise listed).

## Data Availability

All used data are publicly available through the https://ida.loni.usc.edu/ website.

## Funding

This work was supported by Row Fogo Charitable Trust (grant no. BRO-D.FID3668413, MVH). This study was supported by grants from the German Research Foundation (SCHR 774/5-1 to MLS), and the eHealthSax Initiative of the Sächsische Aufbaubank (Project TelDem). Accordingly, this study was co-financed with tax revenue based on the budget approved by the Saxon state parliament.

Data collection and sharing for this project was funded by the Alzheimer’s Disease Neuroimaging Initiative (ADNI) (National Institutes of Health Grant U19 AG024904) and DOD ADNI (Department of Defense award number W81XWH-12-2-0012). ADNI is funded by the National Institute on Aging, the National Institute of Biomedical Imaging and Bioengineering, and through generous contributions from the following: AbbVie, Alzheimer’s Association; Alzheimer’s Drug Discovery Foundation; Araclon Biotech; BioClinica, Inc.; Biogen; Bristol-Myers Squibb Company; CereSpir, Inc.; Cogstate; Eisai Inc.; Elan Pharmaceuticals, Inc.; Eli Lilly and Company; EuroImmun; F. Hoffmann-La Roche Ltd and its affiliated company Genentech, Inc.; Fujirebio; GE Healthcare; IXICO Ltd.; Janssen Alzheimer Immunotherapy Research & Development, LLC.; Johnson & Johnson Pharmaceutical Research & Development LLC.; Lumosity; Lundbeck; Merck & Co., Inc.; Meso Scale Diagnostics, LLC.; NeuroRx Research; Neurotrack Technologies; Novartis Pharmaceuticals Corporation; Pfizer Inc.; Piramal Imaging; Servier; Takeda Pharmaceutical Company; and Transition Therapeutics. The Canadian Institutes of Health Research is providing funds to support ADNI clinical sites in Canada. Private sector contributions are facilitated by the Foundation for the National Institutes of Health (www.fnih.org). The grantee organization is the Northern California Institute for Research and Education, and the study is coordinated by the Alzheimer’s Therapeutic Research Institute at the University of Southern California. ADNI data are disseminated by the Laboratory for Neuro Imaging at the University of Southern California.

## Author contributions

Conceptualization: G.G., J.R, K.T, M.L.S

Methodology: G.G., J.R, P.G.M., P.E., K.T, M.L.S

Investigation: G.G., J.R, P.E., K.T, M.L.S

Visualization: G.G., K.T., N.S., E.N.M.

Supervision: J.R, K.T., E.N.M., M.L.S

Writing—original draft: G.G., J.R., E.N.M.

Writing—review & editing: All authors

## Competing interests

All authors declare they have no competing interests.

## Supplementary Materials

### 1. Figures

**Supplementary Figure 1.**
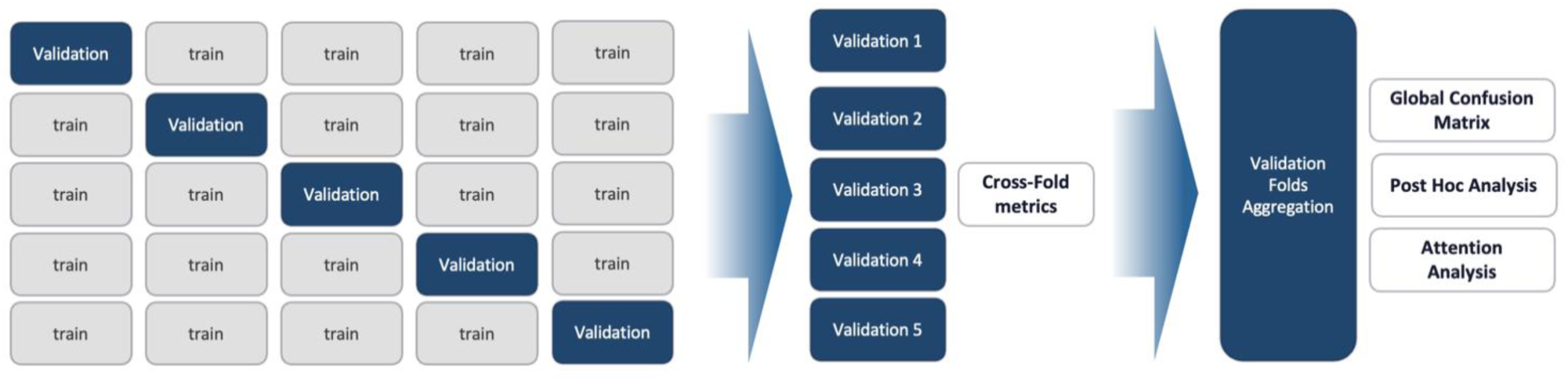
Description of the 5-Fold valuation process. Five models were trained on different data from different participants in a k-fold cross-validation setup. Then average accuracy and standard deviations were collected from the different models on the respective validation folds. Finally, explainability metrics and post hoc analyzes were conducted on each validation fold and aggregated to provide a dataset-level analysis.

**Supplementary Figure 2.**
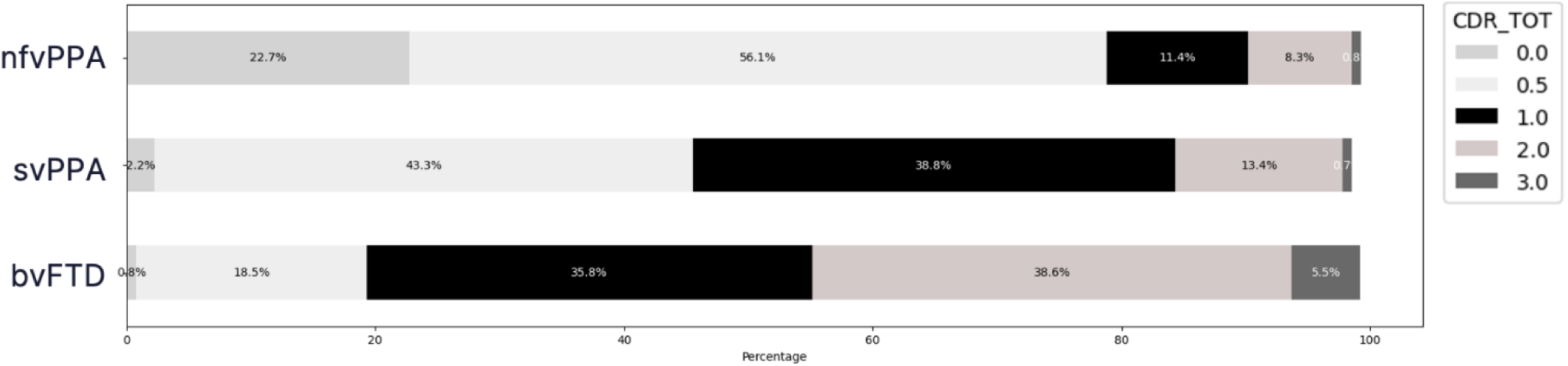
Distribution of CDR Scores in FTLDNI Dataset. We used the distributions described in the original CDR plus NACC FTLD as a reference and compared this to our input CDR values. Given the similarity between the original version and the version used in our modeling, we conclude that the score matches the classic formulation of the CDR global score, thereby reducing the chances of biasing our model.

### 2. Tables

**Supplementary Table1:** Characteristics and distribution of endpoints of studies’ participants across datasets.

|  |  |  |  |
| --- | --- | --- | --- |
|  | **ADNI**<br>**(n=9138)** | **AIBL**<br>**(n=1689)** | **FTLDNI**<br>**(n=1106)** |
| Age | 74.75 ± 7.46<br>missing: 0% | 75.93 ± 6.76<br>missing: 0.1% | 64.83 ± 7.41<br>missing: 0% |
| Sex | 'F': N=4961,<br>'M': N=4176<br>missing: 0% | 'F': N=876,<br>'M': N=813<br>missing: 0% | 'F': N=532,<br>'M': N=574,<br>missing: 0% |
| Years of education | 16.04 ± 2.75<br>missing: 0% | — | 16.52 ± 2.51<br>missing: 2.4% |
| MCI conversion time<br>(months) | 27.71 ± 24.44 | 25.42 ± 11.45 | — |
| MMSE<br>(total score) | 26.84 ± 3.76<br>missing: 6.8% | 27.47 ± 3.75<br>missing: 0.2% | 25.73 ± 5.90<br>missing 13.7% |
| MoCA<br>(total score) | 23.86 ± 4.46<br>missing: 83.0% | — | 25.73 ± 5.90<br>missing: 54.2% |
| Geriatric Depression Scale<br>(total score) | 1.61 ± 1.86<br>missing: 18.7% | — | 2.73 ± 3.24<br>missing: 25.5% |
| CDR<br>(global score) | 0.42 ± 0.42<br>missing: 7.5% | 0.21 ± 0.41<br>missing: 0.4% | 0.21 ± 0.41<br>missing: 20.2% |
| WMS IV - Logic Memory II<br>(Total Number of Story<br>Units Recalled) | 8.60 ± 6.19<br>missing: 24.7% | 9.41 ± 5.39<br>missing: 1% | — |
| WMS IV - Logic Memory I<br>(Total Number of Story<br>Units Recalled) | 10.59 ± 5.53<br>missing: 24.5% | 10.98 ± 4.87<br>missing: 1.4% | — |
| APOE | (3,3): N=4330,<br>(3,4): N=2909,<br>(4,4): N=821,<br>(2,3): N=685,<br>(2,4): N=192,<br>(2,2): N=20<br>missing: 2.0% | (3,3): N=850,<br>(3,4): N=469,<br>(4,4): N=201,<br>(2,3): N=98,<br>(2,4): N=40,<br>(2,2): N=4<br>missing: 1.6% | — |
| Amyloid-$\beta_{1-42}$ (CSF) | 912.03 ± 439.52<br>missing: 73.5% | — | — |
| Amyloid-$\beta_{1-40}$ (CSF) | 16813.19 ± 4507.24<br>missing: 93.0% | — | — |
| Tau (CSF) | 256.69 ± 84.97<br>missing: 73.7% | — | — |
| P-Tau (CSF) | 23.94 ± 9.09<br>missing: 73.7% | — | — |
| NFL (CSF) | 1466.19 ± 1036.36<br>missing 95% | — | — |
| Gap43 (Plasma) | 4989.20 ± 2218.84<br>missing: 90.2% | — | — |
| NT1 Tau (Plasma) | $2.28 \pm 0.60$<br>missing: 94.4% | — | — |
| pTau<sub>181</sub> (Plasma) | $15.59 \pm 7.21$<br>missing 76.4% | — | — |
| NFL (Plasma) | $36.43 \pm 13.54$<br>missing 76.3% | — | — |
| T1w MRI | N=9124<br>missing: 0.15% | N=1118<br>missing: 33.8% | N=1039<br>missing: 6.06% |

**Supplementary Table 2:** Characteristics and distribution of endpoints of studies’ participants across diagnoses.

|  |  |  |  |  |  |  |  |
| --- | --- | --- | --- | --- | --- | --- | --- |
|  | **AD**<br>**(n=1931)** | **CN**<br>**(n=4781)** | **sMCI**<br>**(n=2767)** | **pMCI**<br>**(n=1030)** | **bvFTD**<br>**(n=264)** | **nfvPPA**<br>**(n=137)** | **svPPA**<br>**(n=147)** |
| Age | 75.78 ± 7.64<br>missing: 0% | 74.15 ± 7.38<br>missing: 0% | 74.39 ± 7.94<br>missing: 0% | 74.52 ± 7.49<br>missing: 0% | 62.53 ± 5.78<br>missing: 0% | 68.26 ± 7.39<br>missing: 0% | 63.59 ± 6.20<br>missing: 0% |
| Sex | 'F':<br>N=1072,<br>'M':<br>N=859<br>missing: 0% | 'F':<br>N=2396,<br>'M':<br>N=2385<br>missing: 0% | 'F':<br>N=1554,<br>'M':<br>N=1212,<br>missing: 0% | 'F':<br>N=616,<br>'M':<br>N=414,<br>missing: 0% | 'F':<br>N=90,<br>'M':<br>N=174<br>missing: 0% | 'F':<br>N=74,<br>'M':<br>N=63<br>missing: 0% | 'F':<br>N=62,<br>'M':<br>N=85<br>missing: 0% |
| Years of education | 15.46 ± 2.86<br>missing: 8.2% | 16.61 ± 2.47<br>missing: 26.7% | 16.0 ± 2.83<br>missing: 7.0% | 15.89 ± 2.64<br>missing: 6.2% | 15.85 ± 2.86<br>missing: 1.9% | 16.05 ± 2.64<br>missing: 1.5% | 16.44 ± 2.64<br>missing: 2.0% |
| MMSE (total score) | 21.58 ± 4.56<br>missing: 4.3% | 29.00 ± 1.24<br>missing: 4.7% | 27.76 ± 2.12<br>missing: 8.8% | 26.26 ± 2.63<br>missing: 7.3% | 22.34 ± 6.80<br>missing: 9.5% | 24.57 ± 5.63<br>missing: 19.7% | 22.17 ± 6.86<br>missing: 16.3% |
| MoCA (total score) | 16.36 ± 5.21<br>missing: 92.4% | 26.47 ± 2.54<br>missing: 78.8% | 22.98 ± 3.35<br>missing: 84.9% | 20.66 ± 2.88<br>missing: 96.6% | 16.84 ± 7.33<br>missing: 47.3% | 19.67 ± 6.45<br>missing: 57.7% | 16.49 ± 5.76<br>missing: 53.7% |
| Geriatric Depression Scale (total score) | 1.99 ± 2.04<br>missing: 27.2% | 1.03 ± 1.51<br>missing: 36.3% | 1.85 ± 1.84<br>missing: 24.6% | 1.81 ± 1.79<br>missing: 35.7% | 4.05 ± 3.60<br>missing: 26.1% | 2.96 ± 3.03<br>missing: 28.5% | 4.85 ± 3.79<br>missing: 36.1% |
| CDR<br>(global<br>score) | 0.95 ±<br>0.49<br>missing:<br>4.8% | 0.03 ±<br>0.13<br>missing:<br>7.5% | 0.46 ±<br>0.16<br>missing:<br>9.5% | 0.53 ±<br>0.17<br>missing:<br>7.3% | 1.40 ±<br>0.71<br>missing:<br>4.5% | 0.59 ±<br>0.56<br>missing:<br>4.4% | 0.91 ±<br>0.54<br>missing:<br>10.2% |
| WMS IV -<br>Logic<br>Memory II<br>(Total<br>Number of<br>Story<br>Units<br>Recalled) | 1.35 ±<br>2.54<br>missing:<br>24.7% | 13.03 ±<br>4.13<br>missing:<br>20.5% | 7.90 ±<br>4.79<br>missing:<br>26.2% | 3.79 ±<br>3.67<br>missing:<br>35.5 | — | — | — |
| WMS IV -<br>Logic<br>Memory I<br>(Total<br>Number of<br>Story<br>Units<br>Recalled) | 4.03 ±<br>3.29<br>missing:<br>23.9% | 14.20 ±<br>3.84<br>missing:<br>20.4% | 10.16 ±<br>4.31<br>missing:<br>26.1% | 7.33 ±<br>3.61<br>missing:<br>35.4% | — | — | — |
| APOE | (3,3):<br>N=559,<br>(3,4):<br>N=859,<br>(4,4):<br>N=384,<br>(2,3):<br>N=47,<br>(2,4):<br>N=51,<br>(2,2):<br>N=4<br>missing:<br>1.4% | (3,3):<br>N=2428,<br>(3,4):<br>N=1054,<br>(4,4):<br>N=112,<br>(2,3):<br>N=546,<br>(2,4):<br>N=85,<br>(2,2):<br>N=19<br>missing:<br>1.6% | (3,3):<br>N=1435,<br>(3,4):<br>N=807,<br>(4,4):<br>N=198,<br>(2,3):<br>N=203,<br>(2,4):<br>N=51,<br>(2,2): N=1<br>missing:<br>2.6% | (3,3):<br>N=340,<br>(3,4):<br>N=469,<br>(4,4):<br>N=164,<br>(2,3):<br>N=24,<br>(2,4):<br>N=33,<br>(2,2):<br>N=0<br>missing:<br>0% | — | — | — |
| Amyloid-$\beta_{1-42}$ (CSF) | 613.89 ± 257.11<br>missing: 76.2% | 1047.08 ± 391.32<br>missing: 84.4% | 908.02 ± 391.09<br>missing: 80.2% | 678.80 ± 255.25<br>missing: 73.8% | — | — | — |
| Amyloid-$\beta_{1-40}$ (CSF) | 16770.82 ± 4764.81<br>missing: 97.2% | 16607.53 ± 4078.15<br>missing: 92.7% | 16270.73 ± 4568.60<br>missing: 95.8% | 17579.74 ± 4020.97<br>missing: 98.2% | — | — | — |
| Tau (CSF) | 295.38 ± 69.69<br>missing: 82.9% | 229.09 ± 70.18<br>missing: 81.7% | 237.76 ± 74.38<br>missing: 79.0% | 276.22 ± 74.37<br>missing: 80.5% | — | — | — |
| P-Tau (CSF) | 28.40 ± 7.60<br>missing: 82.7% | 20.54 ± 6.96<br>missing: 81.8% | 22.04 ± 7.86<br>missing: 78.9% | 26.63 ± 8.11<br>missing: 80.5% | — | — | — |
| NFL (CSF) | 1375.24 ± 344.55<br>missing 95.9% | 1057.09 ± 377.31<br>missing 97.9% | 1269.81 ± 383.57<br>missing 97.6% | 1300.45 ± 363.73<br>missing 92.5% | — | — | — |
| Gap43 (Plasma) | 5560.02 ± 1881.73<br>missing: 94.5% | 4596.43 ± 1865.51<br>missing: 94.1% | 4436.70 ± 1895.26<br>missing: 88.8% | 5353.33 ± 1768.72<br>missing: 90.9% | — | — | — |
| NT1 Tau<br>(Plasma) | 2.31 ±<br>0.54<br>missing:<br>94.7% | 2.24 ±<br>0.64<br>missing:<br>94.9% | 2.31 ±<br>0.54<br>missing:<br>96.4% | 2.38 ±<br>0.60<br>missing:<br>95.8% | — | — | — |
| pTau<sub>181</sub><br>(Plasma) | 19.50 ±<br>5.73<br>missing<br>85.4% | 13.28 ±<br>6.12<br>missing<br>85.1% | 14.20 ±<br>6.45<br>missing<br>72.8% | 16.85 ±<br>6.01<br>missing<br>84.4% | — | — | — |
| NFL<br>(Plasma) | 41.89 ±<br>5.73<br>missing<br>76.3% | 33.49 ±<br>11.66<br>missing<br>85.1% | 32.99 ±<br>12.06<br>missing<br>72.9% | 37.57 ±<br>10.50<br>missing:<br>84.3% | — | — | — |
| T1w MRI | N=1873<br>missing:<br>3% | N=4332<br>missing:<br>9.39% | N=2701<br>missing:<br>2.39% | N=1005<br>missing:<br>2.43% | N=247<br>missing:<br>6.44% | N=122<br>missing:<br>10.95% | N=136<br>missing:<br>7.48% |

### 3. State-of-the-art (SOTA) - Comparisons to existing literature

To compare our results to existing literature, we collected the balanced accuracies reported by the below studies. When the balanced accuracy score was not provided, we relied on sensitivity and specificity to compute the metric by summing the sensitivity and specificity metrics and dividing number that by 2.

## Notes

### Competing Interest Statement

The authors have declared no competing interest.

### Author Declarations

All used data are publicly available through the https://ida.loni.usc.edu/ website.

### Summary of Updates

Correction of typos, correction of duplicated informatin in Figure 7, improved resolution of figures.

